# RF power, B_1+rms_, and SAR variation with RF coils, conductive metallic implants, and ionic solutions at 1.5T and 3T

**DOI:** 10.64898/2026.01.04.26343414

**Authors:** David H. Gultekin

## Abstract

**Background and Purpose:** The magnetic resonance imaging (MRI) access for patients with active and passive implants is limited by radiofrequency (RF) safety. The time-averaged root-mean-square RF field (B_1+rms_) and specific absorption rate (SAR) are being evaluated to monitor and control RF-induced heating near conductive metallic implants, such as deep brain stimulation (DBS) leads, during MRI. However, experimental methods to assess the relationship between RF power, B_1+rms,_ and SAR are lacking for RF coils, metallic implants, and ionic solutions.

**Materials and Methods:** A method is developed to evaluate the variation of RF power, B_1+rms,_ and SAR with RF coils, metallic implants, and ionic solutions using phantoms consisting of water (H_2_O) and sodium chloride (NaCl) with four ionic concentrations (0, 1, 2, 3 %), four metallic wavelengths (0, *λ*/2, *λ*, 2*λ*), two RF coils (body, head) transmit/receive (Tx/Rx) combinations, and five RF pulse flip angles (30°, 45°, 60°, 75°, 90°) in two B_0_ fields (1.5T and 3T).

**Results:** The scanner-reported RF power and SAR varied with RF pulse sequences, RF coils, Tx/Rx, metallic implants, and ionic solutions, whereas B_1+rms_ varied only with RF pulse sequences. The RF power, B_1+rms_, and SAR relationship depends on RF pulse sequences, RF coils, Tx/Rx, implant wavelengths, and ionic concentrations. SAR (whole-body, head) scaled with RF power by absorption ratios (α) variable with experimental conditions.

**Conclusions:** B_1+rms_ is insensitive to the presence and absence of conductive metallic implants and ionic solutions, implant wavelengths, ionic concentrations, RF coils, and Tx/Rx combinations. RF power must be monitored because scanner-reported SAR may vary unpredictably with experiments.

## 1. INTRODUCTION

There is a growing number of patients with conductive metallic implants and electronic devices that require magnetic resonance imaging (MRI) for diagnosis and therapy response monitoring. More than 4.6 million hip and knee procedures, including primary and revision arthroplasty procedures, have been performed between 2012 and 2024 [1]. More than 400,000 patients in the US and about 1.7 million patients in the world receive cardiac implantable electronic devices (CIED) every year [2, 3]. Up until now, it is estimated that more than 300,000 patients have received deep-brain stimulation (DBS) leads globally, and the number is increasing with time at a rate of about 12,000 per year [4]. The applications of electrical stimulation in neuroscience and neurosurgery have led to important advances in surgical treatments of neurological diseases and continue to grow in a wide range of neurological conditions [5–13].

However, conductive metallic implants may heat up significantly during the MRI and create safety risks for the patients [14–18]. The time-varying electromagnetic fields in the MRI may induce heating in the leads and potentially damage the cells in the vicinity of the leads implanted in the tissues [19–23]. Therefore, it is important to develop safety models and methods to predict and control the magnitude of heating in the conductive implants during the MRI [24–26].

In general, most adverse effects reported about the MRI involve electromagnetic power deposition, causing thermal damage to the patients [27, 28]. As a consequence, the potential applications of MRI are limited partly by considerations of radiofrequency (RF) safety that impose constraints on the design of RF coils and RF pulse sequences [29]. A current dilemma in the MRI is how to optimize image quality and scan time while maintaining RF safety. Often, one factor may be improved at the expense of other factors.

There is a growing need and interest in assessing the utility of temperature rise (dT), specific absorption rate (SAR), and the time-averaged root mean square RF fields (B_1+rms_) to monitor RF absorption and thermal hazards in tissues, especially in the presence of conductive metallic implants such as DBS electrodes [30–34]. However, it is not clear how these safety measures correlate with each other as well as with heating and thermal damage to the tissues exposed to RF power by RF coils and pulse sequences [35]. Monitoring SAR and B_1+rms_ by themselves do not include the exposure time, whereas monitoring dT includes the exposure time and the transient effects of RF power absorption. Therefore, measurement of the transient temperature during MRI can help monitor RF safety when RF power and B_1+rms_ remain constant during the scan.

In a previous study, the relationship between RF-induced heating and temperature rise near a conductive metallic DBS lead, B_1+rms_, and RF power for RF pulse sequences, RF coils, and transmit and receive (Tx/Rx) combinations was investigated [35]. It was demonstrated that both temperature rise and RF power varied with RF pulse sequences, RF coils, and Tx/Rx combinations, whereas B_1+rms_ varied only with RF pulse sequences but not with RF coils and Tx/Rx combinations. The RF induced heating and temperature rise were associated with RF power propagation and loss corresponding to the B_1+rms_ for RF coils and Tx/Rx combinations.

However, transient temperature measurements take a much longer time as the temperature increases during the heating period over the scan duration and decreases during the cooling period, which may take several minutes of waiting time between the experiments. There is also a global heating of the medium in a finite-size phantom with repeated experiments, which may require hours of cooling time to return to baseline after several RF-intensive experiments. During the MRI, temperature changes (increases when RF pulses are on and decreases when RF pulses are off), while RF power and B_1+rms_ stay constant.

In this study, the focus is on the relationship between the scanner-reported RF power, B_1+rms_, and SAR varying with RF pulse sequences, RF coils, Tx/Rx combination, conductive metallic implant wavelengths, and conductive ionic concentrations. This method can be used to compare the results of the experimental measurements to the computational electromagnetic modeling and thermal modeling to design and optimize RF coils and RF pulse sequences for RF safety and RF-induced heating near the conductive metallic implants during the MRI.

## 2. METHODS AND MATERIALS

### 2.1 Theory

In nuclear magnetic resonance imaging, nuclear spins are placed in a static magnetic field (*B_o_*) and perturbed away from the equilibrium magnetization by an external transverse magnetic field (*B_1_*), and the resultant signal is received and processed spatially and temporally using the sets of gradient coils and radiofrequency coils. The relationship between the nuclear spin phase (θ) and the magnetic field (*B_1_*) can be derived from Larmor’s equation.

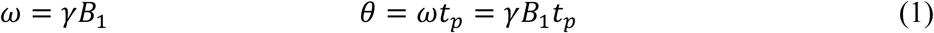

where ω is the angular Larmor frequency (rads/s), γ is the angular gyromagnetic ratio (rads/s·T), *B_1_* is an external transverse magnetic field (Tesla), and *t_p_* is the pulse length (s). A plot of *B_1_* vs. θ yields a straight line with a slope proportional to 1/γ *t_p,_* which can be measured experimentally.

The specific absorption rate in an ex vivo tissue or a substance in the absence of perfusion can be related to the electric field or the initial rate of temperature rise at a point in the body, as

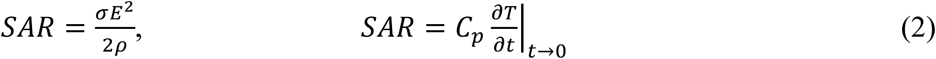

where σ is the electrical conductivity (S/m), *E* is the root-mean-square (rms) electric field (V/m), ρ is the mass density (kg/m^3^), *C_p_* is the specific heat (J/kg·K) in tissue, and (∂*T*/∂*t*)_*t*→0_ is the initial temperature rise with time (K/s) [36].

The whole-body average SAR can be related to the average incident RF power (*P*) as

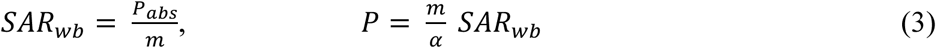

where *m* is the patient’s weight (kg) and *α* = *P_abs_*/*P* is a dimensionless parameter called the absorption ratio, the ratio of the absorbed power (*P_abs_*) to the incident power (*P*). A linear regression of *P* vs. SAR_wb_ yields a straight line with a slope proportional to *m/α,* which can be measured experimentally.

In a simplest coil element, the *E* field magnitude and the incident RF power (P) in a loop of wire with a radius (r) can be formulated using Eqs. (2) and (3) as

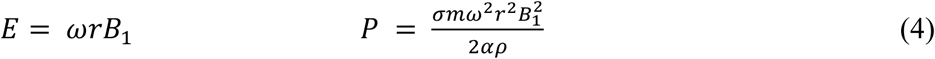

where ω is the angular frequency, *B_1_* is the magnitude of the RF field [37–39]. A linear regression of *P* vs. the square of *B_1_* field yields a straight line with a slope proportional to *σmω*^2^*r*^2^/2*αρ*, which can be measured experimentally.

Further, by substituting Eq. (1) into Eq. (4) and scaling it with the dimensionless parameters, absorption ratio and duty factor, the absorbed RF power and incident RF power can be related to the flip angle (θ) as

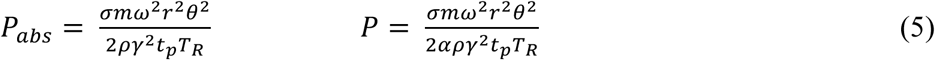

where *α* = *P_abs_*/*P* and *D* = *t_p_*/*T_R_* is the ratio of pulse length (*t_p_*) to the repetition time (T_R_) [37–39]. The exact relation for power deposition depends on the direction of the B_1_ field and the coordinate system in the tissue [27, 40].

RF coils for MRI require a complicated electromagnetic design and optimization. However, the simplest element of a loop and a capacitor (LC) still makes up the unit element of arrays with up to 128 LC loops [29]. The relationship between the RF power, magnetic field, and electric field in a coil depends on a specific coil design and a number of parameters [29, 41, 42]. The optimization of these parameters in an RF coil design is complex and requires electromagnetic modeling and experimentation. However, the RF power and B_1_ field can be monitored for a coil or a combination of coils experimentally. Here, the variation in RF power, B_1+rms_, and SAR is evaluated in various experimental conditions by varying the conductive metallic wavelengths and ionic concentrations using the commercially available RF coils and RF pulse sequences in two MR systems.

### 2.2 MRI Systems and RF Coils

Two MRI systems (Magnetom, 1.5T Aera and 3T Prisma, both Syngo MR XA30A, Siemens Healthcare GmbH, Erlangen, Germany) were used in this study. In a previous study, five RF coil combinations, two Tx/Rx volume coils, a circularly polarized head coil (CP-H) and a body coil (BC), and three Rx only phased-array surface coils, a 20-channel head and neck coil (20-H/N), a 32-channel head coil (32-H), and a 64-channel head and neck coil (64-H/N) were evaluated at 3T Prisma [35]. In this study, a widely used combination of RF coils, a BC Tx/Rx, and a 20-H/N Rx, in two Tx/Rx combinations were used at both 1.5T and 3T to reduce the number of experiments. When the 20-H/N coil was turned off, the Tx/Rx was BC/BC, and when the 20-H/N coil was turned on, the Tx/Rx was BC/20-H/N. The BC specifications (maximum applied B_1+_ and B_1+rms_) were 27 μT and 6.2 μT for 1.5T, and 30 μT and 3.6 μT for 3T. The BC Tx field attenuation (3 dB and 10 dB) from the isocenter was 0.18 m and 0.38 m for 1.5T, and 0.15 m and 0.25 m for 3T.

### 2.3 Aqueous Solutions of NaCl–H2O

Aqueous solutions of sodium chloride (NaCl) and water (H_2_O) with four concentrations (C) of 0, 1, 2 and 3% NaCl by weight corresponding to molar concentrations of 0, 171, 342, and 513 mM (millimole per liter), and specific gravities (SG) of 1.000, 1.005, 1.013, 1.020 kg/L (kilogram per liter), respectively. These electrolytes with monovalent ions also correspond to 0, 171, 342, and 513 mEq/L (milliequivalent per liter) ions of Na^+^ (cations) and Cl^-^ (anions). The NaCl has a molecular weight of 58.44 g/mol (22.99 g/mol Na + 35.45 g/mol Cl) and a maximum solubility of 26.5% by weight or 4.53 M (mol/L) in H_2_O. The electrical conductivity (σ) values for the 0, 1, 2, and 3% NaCl-H_2_O solutions were calculated as 0.00005, 17.8, 34.4, and 51.0 mS/cm (milli Siemens per centimeters), respectively, from the NaCl calibration standards (Ricca Chemical Company, Arlington, TX, USA).

For comparison, normal saline is 0.9% NaCl (9 g/L, 154 mM, 154 mEq/L Na^+^ and Cl^-^) and used for treatment of dehydration and fluid imbalance, and the hypertonic saline is 3% NaCl (30 g/L, 513 mM, 513 mEq/L Na^+^ and Cl^-^), and used for treatment of hyponatremic encephalopathy, traumatic brain injury and cerebral edema. The human serum has a normal range of 135 to 145 mEq/L Na^+^ ions.

### 2.4 Phantoms

A phantom consisting of a conductive metallic wire immersed in a conductive ionic solution was used in this study. Since the implant materials, designs, and positions affect RF induced heating, the same implant with different wavelengths was used to reduce the number of experiments [43–46]. Any conductive metallic wire, such as a DBS lead or any other wire, such as a 14-gauge (Ga) wire, can be used in this method. A 14 Ga stranded wire (64 / 0.2 ± 0.008 mm), copper-clad aluminum (CCA), 8% copper cladding, outside diameter (OD) of 1.85 mm, polyvinylchloride (PVC) insulation, average/minimum thickness of 0.425/0.375 mm, OD of 2.7±0.1 mm, and an electrical resistance (R) of 14.1 Ohm/km was used. The electrical conductivity (α) of the CCA wire was calculated from its resistance as 3.5 × 10^7^ S/m versus 5.96 × 10^7^ S/m for copper (Cu) and 3.77 × 10^7^ S/m for aluminum (Al).

The metallic wire wavelengths (*λ*) of 0, *λ*/2, *λ* and 2*λ* corresponding to lengths (L) of 0, 26, 52 and 104 cm (1.5T) and 0, 13, 26 and 52 cm (3T) were used in combinations with the ionic concentrations (C) of 0, 1, 2 and 3% or 0, 171, 342, and 513 mM NaCl-H_2_O solutions. The lengths of wires of 0, 13, 26, 52, and 104 cm correspond to 0, 2.95, 5.9, 10.8, and 21.6 millimoles of Cu, and 0, 24, 48, 96, and 192 millimoles of Al, respectively.

As shown in **Figure 1** for the 3T phantom, the metallic wire wavelengths of *λ*/2 = 13 cm, *λ* = 26 cm, and 2*λ* = 52 cm were attached to the walls of poly methyl methacrylate (PMMA) plastic containers with dimensions of 22.86 cm, 11.125 cm, and 10.49 cm. A volume of 1.5L of each ionic solution with a height of 6.4 cm in the container was used, and the wire was positioned horizontally at 3.2 cm. The wire had a straight I-shape (*λ*/2), a slightly curved C-shape (*λ*), or a U-shape (2*λ*) in the phantom. The same conductive metallic implant and trajectories were used in the phantoms when testing the ionic solutions, RF pulse sequences, and Tx/Rx combinations.

**Figure 1:**
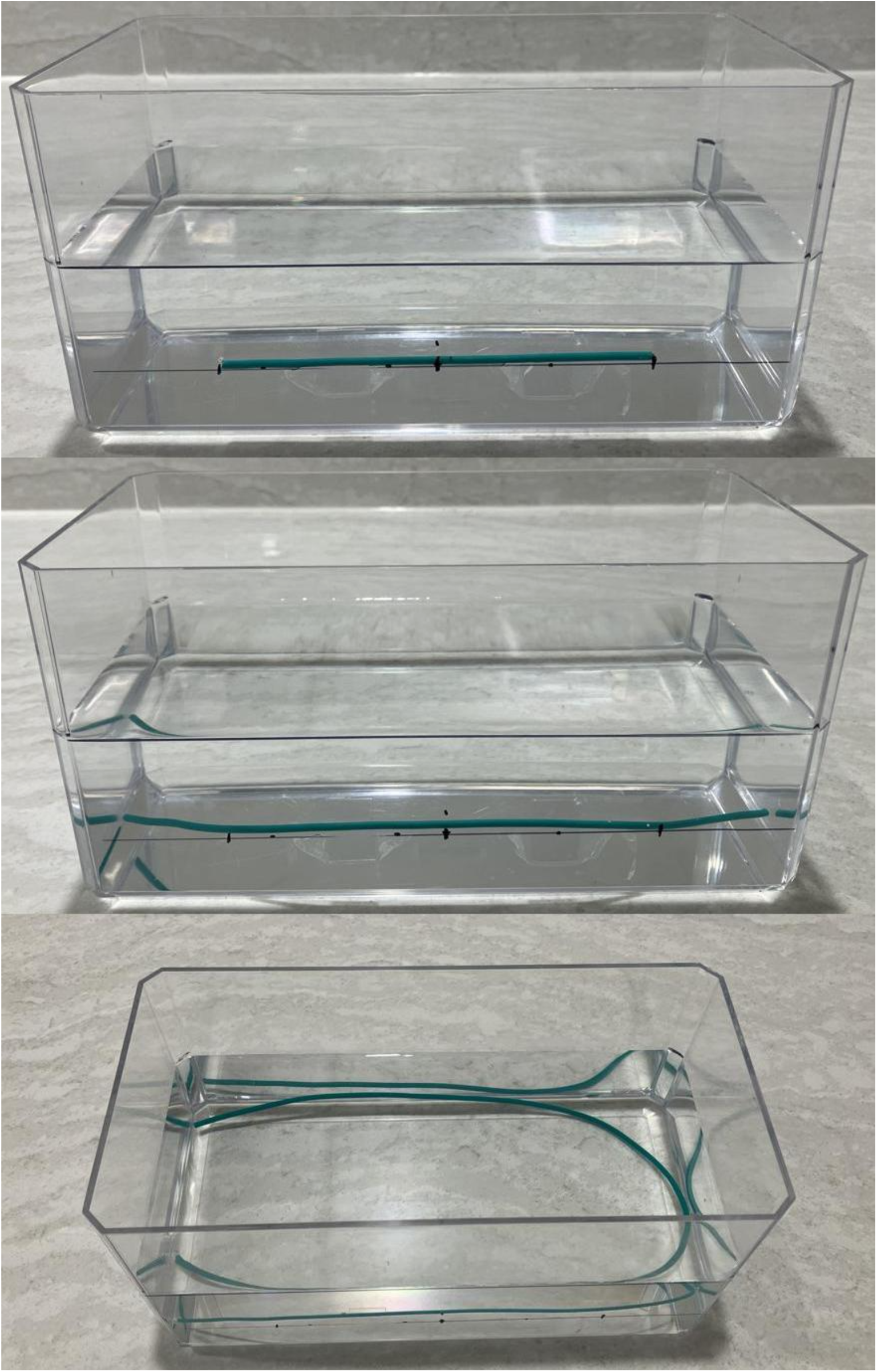
A phantom with conductive ionic solutions of NaCl-H_2_O, conductive metallic wire wavelengths of *λ*/2, *λ*, and 2*λ* corresponding to the conductive metallic wavelengths of 13, 26, and 52 cm, from top to bottom, respectively, at 3T.

### 2.5 RF Pulse Sequences

Five RF pulse sequences were prescribed to vary RF flip angles linearly as an independent parameter. A two-dimensional (2D) gradient echo (GRE) sequence with a variant of the fast low-angle shot (FLASH) sequence was modified with flip angles (θ) of 30, 45, 60, 75, and 90 degrees while keeping all other RF pulse parameters the same. The GRE sequences optimized with nominal parameters were taken from each of the scanners at 1.5T and 3T, respectively. The echo time (T_E_) was 4.76 ms and 2.46 ms, repetition time (T_R_) was 370 ms and 250 ms, slice thickness (z) was 5 mm and 4 mm, the total acquisition time (TA) was 69 and 77 seconds, at 1.5T and 3T, respectively. The number of slices (NS) was 3, the field of view (FOV) was 220 mm × 220 mm, matrix (m) size was 320 × 320. In the nominal sequences, an RF saturation band (SB) of 50 mm parallel to the slice was prescribed at 1.5T, but not at 3T. By default, the RF adjustment strategy was set to standard, B_0_ shimming was tune-up, B_1_ shimming was TrueForm, adjustment tolerance was auto, adjust with body coil was off, confirm frequency was never, and adjustment volume position was set to isocenter [47]. The RF adjustments including B_1_ shimming (available at 3T but not at 1.5T) were performed by the scanner automatically [48]. Also, a previous study showed that RF power and the square of B_1+rms_ scaled linearly with three flip angles (30, 60, 90 degrees) and two RF pulse types (fast and normal), so only one RF pulse type (fast) was prescribed in this study [35].

These five RF sequences were repeated for two RF coil Tx/Rx combinations (BC/BC and BC/20-H/N), four concentrations (C) of aqueous solutions of NaCl-H_2_O (0, 1, 2 and 3%) and four wavelengths (*λ*) of wires (0, *λ*/2, *λ* and 2*λ*) corresponding to four lengths (L) of wires (0, 26, 52 and 104 cm at 1.5T, and 0, 13, 26 and 52 cm at 3T).

The scanner-reported RF power, B_1+rms_, and SAR for the whole body and head (SAR_wb_ and SAR_h_), were recorded with high precision during the experiments. For consistency in SAR computation by the scanner’s algorithm, a constant patient weight, height, and age (68 kg, 1.8 meters, and 20 years) were used for all series. Additionally, transmit reference amplitude (TxRA), transmit frequency (TxF) were recorded, and the absorption ratio (α) was computed from RF power and SAR_wb_ as α = m·SAR_wb_/P for each of the experiments.

### 2.6 Experimental Design and Statistical Analysis

A full factorial design of experiment (DOE) with multiple factors and levels was implemented to study the relationship between the dependent variables (P, B_1+rms_, and SAR) and independent variables (θ, Tx/Rx, L, and C). The experiments consisted of 5 levels of θ (30, 45, 60, 75, and 90 degrees), 2 levels of Tx/Rx combinations (1 and 2), 4 levels of L (0, *λ*/2, *λ* and 2*λ*), and 4 levels of C (0, 1, 2 and 3%), and a total of 160 experiments (5 × 2 × 4 × 4) per B_0_ field and 320 experiments (160 × 2) for both B_0_ fields (1.5T and 3T) with each session spanning over several hours. The linear regression analyses were performed for B_1+rms_ vs. θ, P vs. the square of B_1+rms_, and P vs. SAR_wb_, for Tx/Rx, L, and C, respectively.

A fractional factorial DOE was performed to assess the variation in RF power, B_1+rms_, and SAR with the number of slices, saturation bands, Tx/Rx, and repetition time using a phantom with an aqueous solution of 5g/L NaCl and 3.75 g/L NiSO_4_×6H_2_O at both 1.5T and 3T. Additionally, experiments were performed to assess the effects of saturation band thickness and slice thickness on P, B_1+rms_ and SAR using an RF pulse sequence with three flip angles (30, 60, 90 degrees) and the Tx/Rx combination of BC/20-H/N.

## 3. RESULTS

The scanner-reported time-averaged RF power, time-averaged root-mean-square RF field B_1+rms_, and specific absorption rate for whole-body and head were evaluated versus the flip angles, RF coils Tx/Rx combinations, conductive metallic wavelengths, and ionic concentrations of NaCl-H_2_O solutions.

B_1+rms_ varied linearly with θ per Eq. (1) whereas RF power and SAR varied linearly with the square of θ and square of B_1+rms_ per Eqs. (4) and (5). A linear regression analysis was performed for B_1+rms_ vs. θ, P vs. the square of B_1+rms_, and P vs. SAR_wb_ for 5 RF pulse sequences, and the linear regression parameters (slope and intercept) along with the coefficient of determination (R^2^) were computed for 160 experiments consisting of 32 linear fittings each at 1.5T and 3T, respectively. The slopes of dB_1_/dθ, dP/dB_1_^2^, dP/ΔS_wb,_ and the intercepts of B_1_(0), P(0), and P(0), along with the standard errors, are given in **Tables 1** and **2** for 1.5 and 3T, respectively.

**Table 1:**
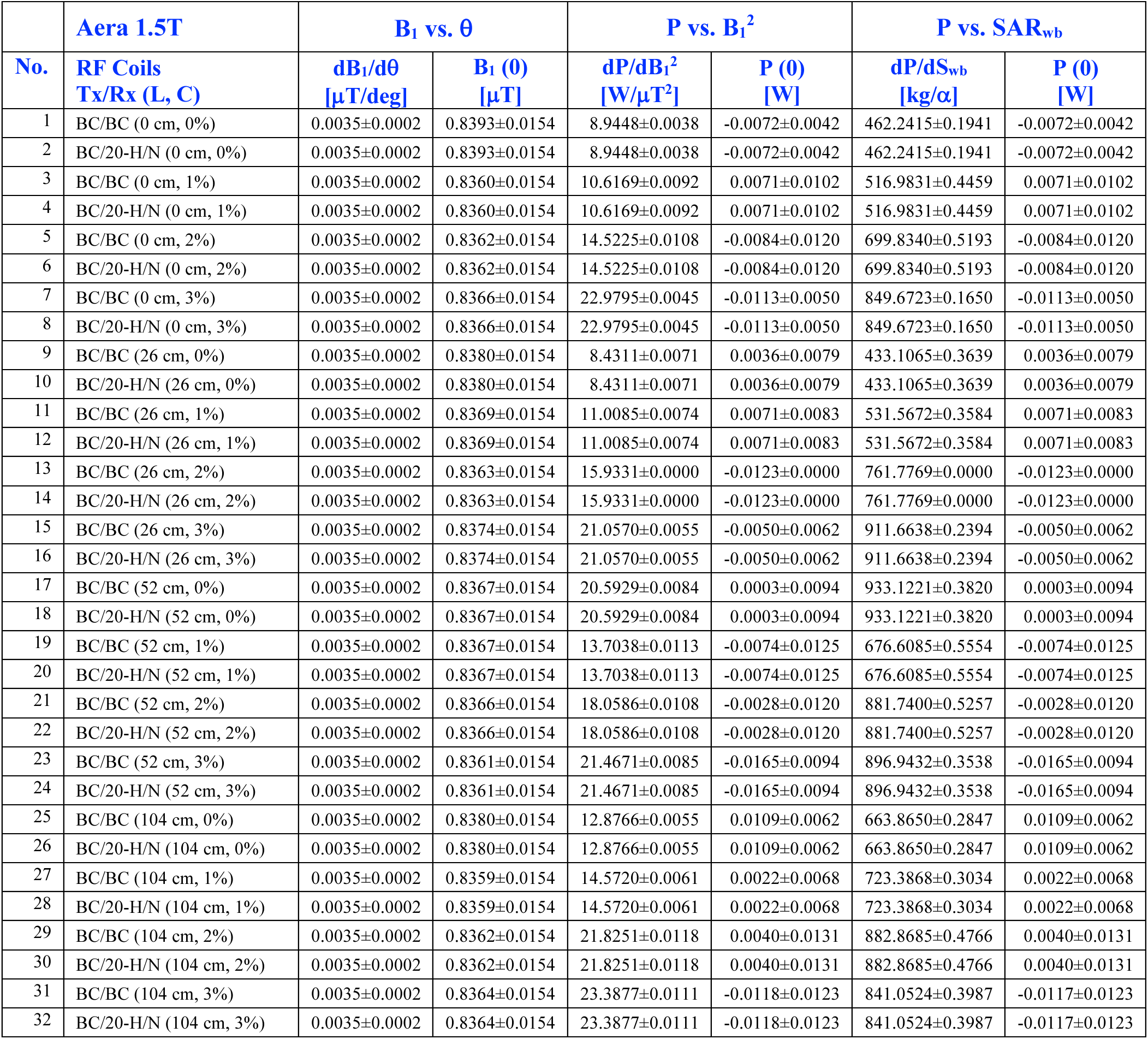
Linear regression results (slope and intercept) for root-mean-square RF field (B_1+rms_) vs. RF pulse flip angle (θ), time-averaged RF power (P) vs. square of B_1+rms_ and whole-body specific absorption rate (SAR_wb_) for RF coils with transmit and receive (Tx/Rx) combinations, conductive metallic implant wavelengths (L), and conductive ionic concentrations (C) of aqueous solutions of sodium chloride (NaCl) and water (H_2_O) at 1.5T.

**Table 2:** Linear regression results (slope and intercept) for root-mean-square RF field (B_1+rms_) vs. RF pulse flip angle (θ), time-averaged RF power (P) vs. square of B_1+rms_ and whole-body specific absorption rate (SAR_wb_) for RF coils with transmit and receive (Tx/Rx) combinations, conductive metallic implant wavelengths (L), and conductive ionic concentrations (C) of aqueous solutions of sodium chloride (NaCl) and water (H_2_O) at 3T.

| | Prisma 3T | $B_1$ vs. $\theta$ | | P vs. $B_1^2$ | | P vs. $SAR_{wb}$ | |
| --- | --- | --- | --- | --- | --- | --- | --- |
| No. | RF Coils<br>Tx/Rx (L, C) | $dB_1/d\theta$<br>[$\mu T/deg$] | $B_1(0)$<br>[$\mu T$] | $dP/dB_1^2$<br>[W/$\mu T^2$] | P (0)<br>[W] | $dP/dS_{wb}$<br>[kg/$\alpha$] | P (0)<br>[W] |
| 1 | BC/BC (0 cm, 0%) | 0.0094±0.0000 | 0.0000±0.0000 | 3.2715±0.0026 | -0.0010±0.0011 | 76.6099±0.0610 | -0.0010±0.0011 |
| 2 | BC/20-H/N (0 cm, 0%) | 0.0094±0.0000 | 0.0000±0.0000 | 3.4138±0.0029 | -0.0021±0.0012 | 79.9424±0.0686 | -0.0021±0.0012 |
| 3 | BC/BC (0 cm, 1%) | 0.0095±0.0000 | 0.0000±0.0000 | 5.5152±0.0038 | 0.0050±0.0016 | 110.5703±0.0759 | 0.0050±0.0016 |
| 4 | BC/20-H/N (0 cm, 1%) | 0.0094±0.0000 | 0.0000±0.0000 | 7.8605±0.0049 | -0.0001±0.0021 | 109.3546±0.0685 | -0.0001±0.0021 |
| 5 | BC/BC (0 cm, 2%) | 0.0095±0.0000 | 0.0000±0.0000 | 7.5683±0.0058 | 0.0043±0.0025 | 110.5145±0.0840 | 0.0043±0.0025 |
| 6 | BC/20-H/N (0 cm, 2%) | 0.0094±0.0000 | 0.0000±0.0000 | 15.9123±0.0079 | -0.0006±0.0034 | 88.5539±0.0441 | -0.0006±0.0034 |
| 7 | BC/BC (0 cm, 3%) | 0.0095±0.0000 | 0.0000±0.0000 | 7.7695±0.0070 | 0.0008±0.0030 | 111.1972±0.1007 | 0.0008±0.0030 |
| 8 | BC/20-H/N (0 cm, 3%) | 0.0094±0.0000 | 0.0000±0.0000 | 19.7675±0.0045 | -0.0040±0.0019 | 75.7073±0.0171 | -0.0040±0.0019 |
| 9 | BC/BC (13 cm, 0%) | 0.0094±0.0000 | 0.0000±0.0000 | 3.0122±0.0000 | 0.0000±0.0000 | 70.5385±0.0000 | 0.0000±0.0000 |
| 10 | BC/20-H/N (13 cm, 0%) | 0.0094±0.0000 | 0.0000±0.0000 | 3.0986±0.0029 | 0.0021±0.0012 | 72.5624±0.0685 | 0.0021±0.0012 |
| 11 | BC/BC (13 cm, 1%) | 0.0095±0.0000 | 0.0000±0.0000 | 5.6690±0.0026 | 0.0010±0.0011 | 110.5300±0.0505 | 0.0010±0.0011 |
| 12 | BC/20-H/N (13 cm, 1%) | 0.0094±0.0000 | 0.0000±0.0000 | 9.3126±0.0055 | -0.0039±0.0024 | 109.1341±0.0649 | -0.0039±0.0024 |
| 13 | BC/BC (13 cm, 2%) | 0.0095±0.0000 | 0.0000±0.0000 | 7.1181±0.0061 | -0.0023±0.0026 | 110.8938±0.0943 | -0.0023±0.0026 |
| 14 | BC/20-H/N (13 cm, 2%) | 0.0094±0.0000 | 0.0000±0.0000 | 17.7112±0.0026 | -0.0010±0.0011 | 81.2992±0.0119 | -0.0010±0.0011 |
| 15 | BC/BC (13 cm, 3%) | 0.0095±0.0000 | 0.0000±0.0000 | 8.6972±0.0055 | -0.0039±0.0024 | 111.0424±0.0697 | -0.0039±0.0024 |
| 16 | BC/20-H/N (13 cm, 3%) | 0.0094±0.0000 | 0.0000±0.0000 | 27.0554±0.0076 | 0.0005±0.0033 | 69.3702±0.0195 | 0.0005±0.0033 |
| 17 | BC/BC (26 cm, 0%) | 0.0094±0.0000 | 0.0000±0.0000 | 1.5187±0.0056 | -0.0029±0.0024 | 35.5636±0.1306 | -0.0029±0.0024 |
| 18 | BC/20-H/N (26 cm, 0%) | 0.0094±0.0000 | 0.0000±0.0000 | 1.2947±0.0026 | -0.0010±0.0011 | 30.3195±0.0618 | -0.0010±0.0011 |
| 19 | BC/BC (26 cm, 1%) | 0.0095±0.0000 | 0.0000±0.0000 | 6.0481±0.0030 | -0.0011±0.0013 | 111.6514±0.0562 | -0.0011±0.0013 |
| 20 | BC/20-H/N (26 cm, 1%) | 0.0095±0.0000 | 0.0000±0.0000 | 12.0841±0.0094 | -0.0002±0.0040 | 107.6392±0.0836 | -0.0002±0.0040 |
| 21 | BC/BC (26 cm, 2%) | 0.0095±0.0000 | 0.0000±0.0000 | 6.8777±0.0061 | 0.0030±0.0027 | 110.9341±0.0989 | 0.0030±0.0027 |
| 22 | BC/20-H/N (26 cm, 2%) | 0.0095±0.0000 | 0.0000±0.0000 | 24.4226±0.0051 | 0.0018±0.0022 | 71.0778±0.0148 | 0.0018±0.0022 |
| 23 | BC/BC (26 cm, 3%) | 0.0095±0.0000 | 0.0000±0.0000 | 6.4879±0.0029 | 0.0021±0.0012 | 111.2169±0.0493 | 0.0021±0.0012 |
| 24 | BC/20-H/N (26 cm, 3%) | 0.0086±0.0005 | 0.0392±0.0320 | 37.1368±0.0061 | -0.0011±0.0024 | 65.2964±0.0106 | -0.0011±0.0024 |
| 25 | BC/BC (52 cm, 0%) | 0.0095±0.0000 | 0.0000±0.0000 | 3.6826±0.0026 | 0.0010±0.0011 | 86.2377±0.0605 | 0.0010±0.0011 |
| 26 | BC/20-H/N (52 cm, 0%) | 0.0094±0.0000 | 0.0000±0.0000 | 4.1841±0.0095 | -0.0002±0.0040 | 97.9804±0.2221 | -0.0002±0.0040 |
| 27 | BC/BC (52 cm, 1%) | 0.0095±0.0000 | 0.0000±0.0000 | 5.9493±0.0055 | -0.0029±0.0024 | 112.2265±0.1040 | -0.0029±0.0024 |
| 28 | BC/20-H/N (52 cm, 1%) | 0.0094±0.0000 | 0.0000±0.0000 | 10.4000±0.0038 | -0.0050±0.0016 | 108.2694±0.0398 | -0.0050±0.0016 |
| 29 | BC/BC (52 cm, 2%) | 0.0095±0.0000 | 0.0000±0.0000 | 7.6349±0.0061 | 0.0023±0.0026 | 111.3332±0.0886 | 0.0023±0.0026 |
| 30 | BC/20-H/N (52 cm, 2%) | 0.0094±0.0000 | 0.0000±0.0000 | 19.6711±0.0039 | -0.0028±0.0017 | 75.7476±0.0150 | -0.0028±0.0017 |
| 31 | BC/BC (52 cm, 3%) | 0.0095±0.0000 | 0.0000±0.0000 | 9.3410±0.0082 | -0.0010±0.0035 | 116.2077±0.1020 | -0.0010±0.0035 |
| 32 | BC/20-H/N (52 cm, 3%) | 0.0095±0.0000 | 0.0000±0.0000 | 23.9288±0.0049 | 0.0001±0.0021 | 71.2071±0.0145 | 0.0001±0.0021 |

The dB_1_/dθ did not change with any of the experiments as a function of Tx/Rx, L, and C, while dP/dB_1_^2^ and dP/dS_w_ changed with each experiment as a function of Tx/Rx, L, and C, as given in **Tables 1** and **2** for 1.5 and 3T, respectively. The difference in dB_1_/dθ, 0.0035 μT/deg vs. 0.0095 μT/deg, in **Tables 1** and **2** was due to the differences in the nominal RF pulse sequences at 1.5T (50 mm saturation band, and T_R_ of 370 ms) and at 3T (no saturation band, and T_R_ of 250 ms), respectively. The intercept of B_1_(0) = 0.83 μT, corresponding to a flip angle of zero degrees, is the B_1+rms_ used for the saturation band in the nominal sequence at 1.5T for all the experiments, as shown in **Table 1**.

B_1+rms_ varied linearly with θ per Eq. (1) independent of B_0_ field, Tx/Rx, L, and C when using the same RF pulse sequences. The number of slices, RF saturation band, and repetition time prescribed in the RF pulse sequences had effects on the B_1+rms_, RF power, and SAR at both 1.5T and 3T. The slope and intercept, along with the R^2^, were computed by fitting B_1_ vs. θ and P vs. B_1_^2^ by varying NS, SB, Tx/Rx, and TR. **Table 3** shows the slope and intercept, dB_1_/dθ and B_1_(0) for B_1_ vs. θ, dP/dB_1_^2^ and P(0) for P vs. B_1_^2^, and the R^2^ for the experiments at 1.5T and 3T, respectively. The dB_1_/dθ was equal at 1.5T and 3T, independent of magnetic field strength (B_0_) per Eq. (1), when SB was off, but not equal when SB was on as shown in **Table 3**.

**Table 3:**
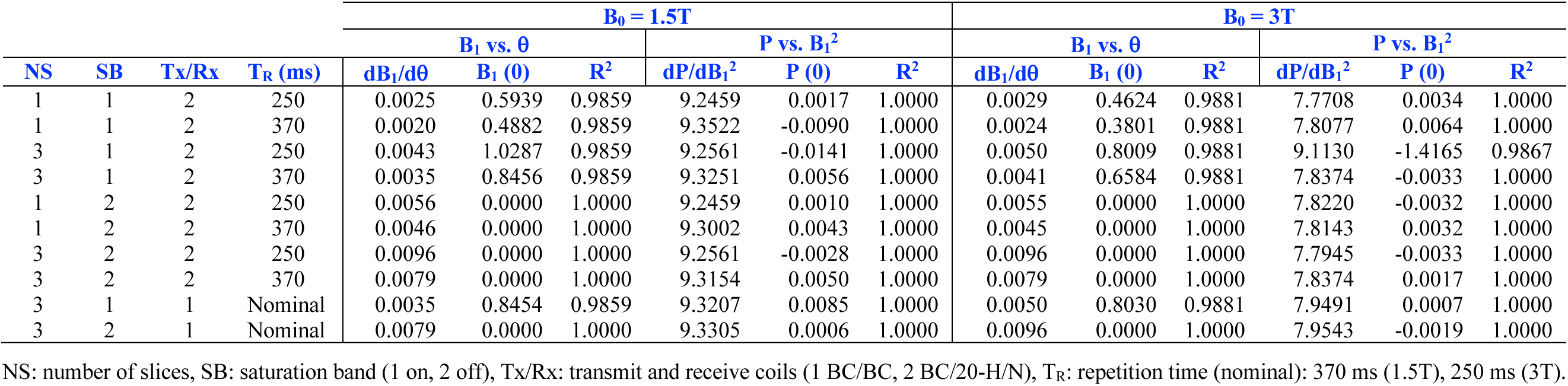
Linear regression results (slope and intercept) for time-averaged root-mean-square RF field (B_1+rms_) vs. RF pulse flip angle (θ), time-averaged RF power (P) vs. square of B_1+rms_ as a function of number of slices (NS), RF saturation bands (SB), RF coils with transmit and receive (Tx/Rx) combinations, and repetition time (T_R_) for an aqueous solution of 5 g/L sodium chloride (NaCl) and 3.75 g/L nickel sulfate (NiSO_4_×6H_2_O) at 1.5T and 3T.

RF power varied linearly with the square of B_1+rms_, consistent with Eqs. (4) and (5), but with different slopes for each of the experiments as a function of Tx/Rx, conductive metallic wavelengths, and ionic concentrations.

The plots of P vs. the square of B_1+rms_ for NaCl-H_2_O solutions with NaCl concentrations of 0% (cyan squares, 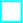), 1% (blue triangles, 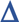), 2% (green diamonds, 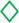) and 3% (red circles, 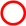) with conductive metallic implant wavelengths of 0*λ*, *λ*/2, *λ* and 2*λ* corresponding to the wavelengths (L) of 0, 26, 52 and 104 cm at 1.5T, and 0, 13, 26 and 52 cm at 3T for Tx/Rx combinations of BC/BC (open legends) and BC/20-H/N (solid legends) with the linear regression lines (dotted lines) and linear regression coefficients (legends) are given in **Figures 2** and **3**, respectively.

**Figure 2:**
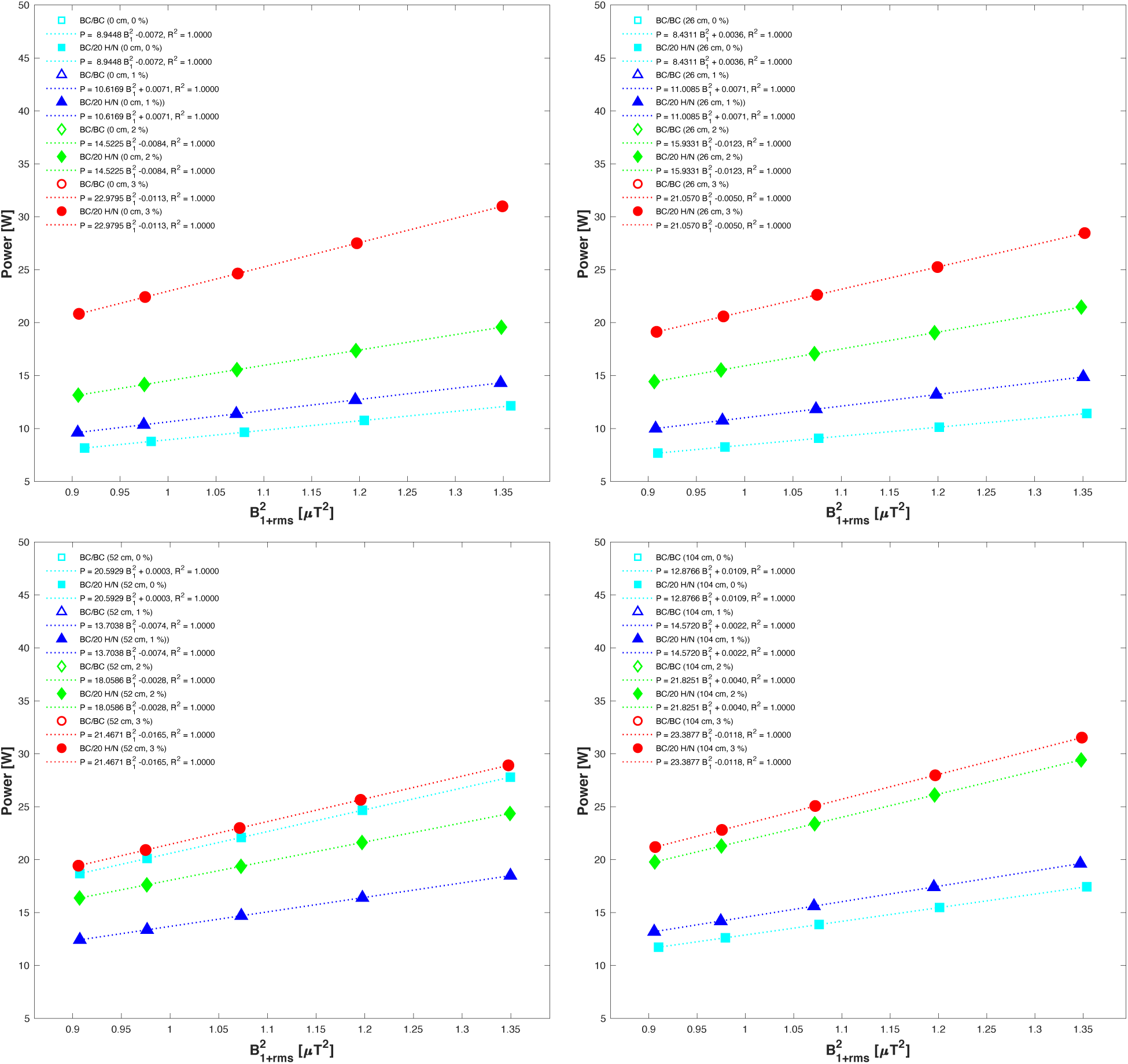
Plots of RF power vs. square of B_1+rms_ for NaCl-H_2_O solutions with four conductive ionic concentrations, 0% (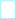), 1% (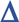), 2% (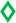) and 3% (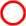), four conductive metallic implant wavelengths, 0, *λ*/2, *λ*, and 2*λ*, corresponding to four wavelengths, 0, 26, 52 and 104 cm, and two Tx/Rx combinations, BC/BC (open legends) and BC/20-H/N (solid legends) at 1.5T.

**Figure 3:**
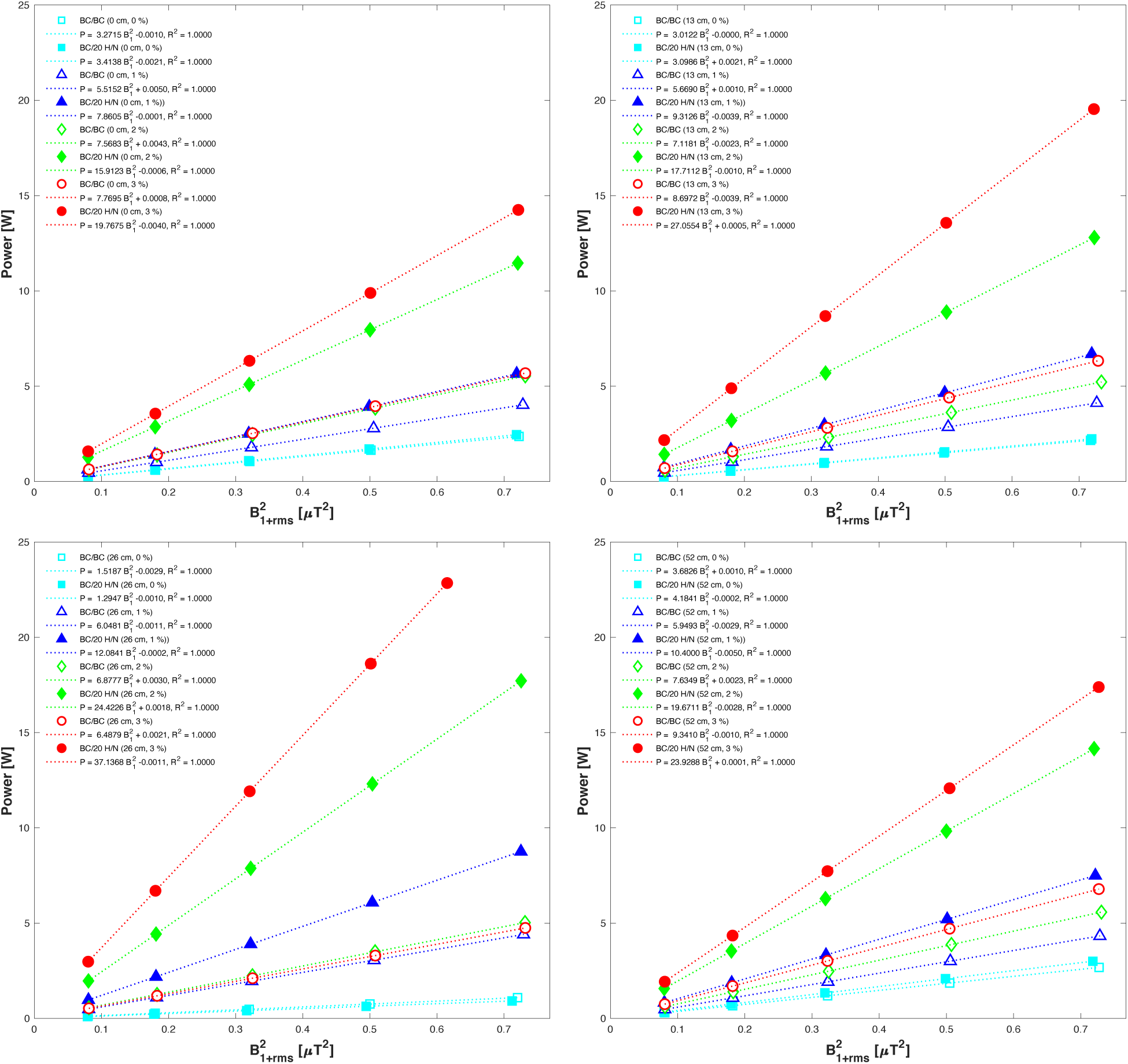
Plots of RF power vs. square of B_1+rms_ for NaCl-H_2_O solutions with four conductive ionic concentrations, 0% (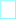), 1% (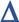), 2% (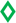) and 3% (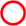), four conductive metallic implant wavelengths, 0, *λ*/2, *λ*, and 2*λ*, corresponding to four wavelengths, 0, 13, 26, and 52 cm, and two Tx/Rx combinations, BC/BC (open legends) and BC/20-H/N (solid legends) at 3T.

RF power varied linearly with whole-body SAR per Eq. (3) but with different absorption ratios for each of the experiments as a function of conductive metallic wavelengths and ionic concentrations.

The plots of P vs. the SAR_wb_ for NaCl-H_2_O solutions with NaCl concentrations of 0% (cyan squares, 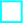), 1% (blue triangles, 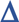), 2% (green diamonds, 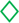) and 3% (red circles, 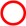) with conductive metallic implant wavelengths of 0*λ*, *λ*/2, *λ* and 2*λ* corresponding to the wavelengths (L) of 0, 26, 52 and 104 cm at 1.5T, and 0, 13, 26 and 52 cm at 3T for Tx/Rx combinations of BC/BC (open legends) and BC/20-H/N (solid legends) with the linear regression lines (dotted lines) and linear regression coefficients (legends) are given in **Figures 4** and **5**, respectively.

**Figure 4:**
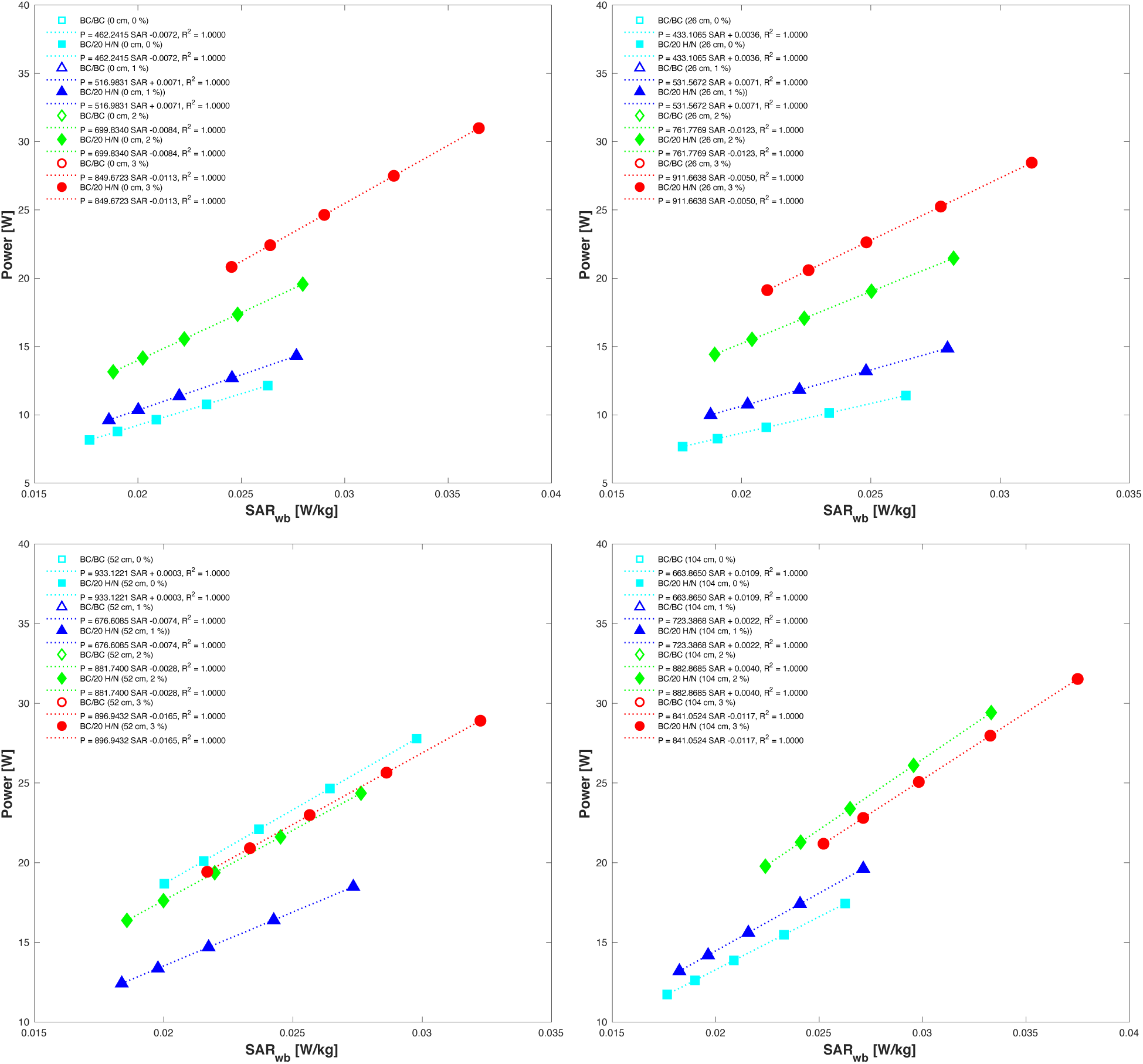
Plots of RF power vs. whole-body SAR for NaCl-H_2_O solutions with four conductive ionic concentrations, 0% (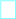), 1% (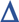), 2% (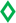) and 3% (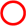), four conductive metallic implant wavelengths, 0, *λ*/2, *λ*, and 2*λ*, corresponding to four wavelengths, 0, 26, 52 and 104 cm, and two Tx/Rx combinations, BC/BC (open legends) and BC/20-H/N (solid legends) at 1.5T.

**Figure 5:**
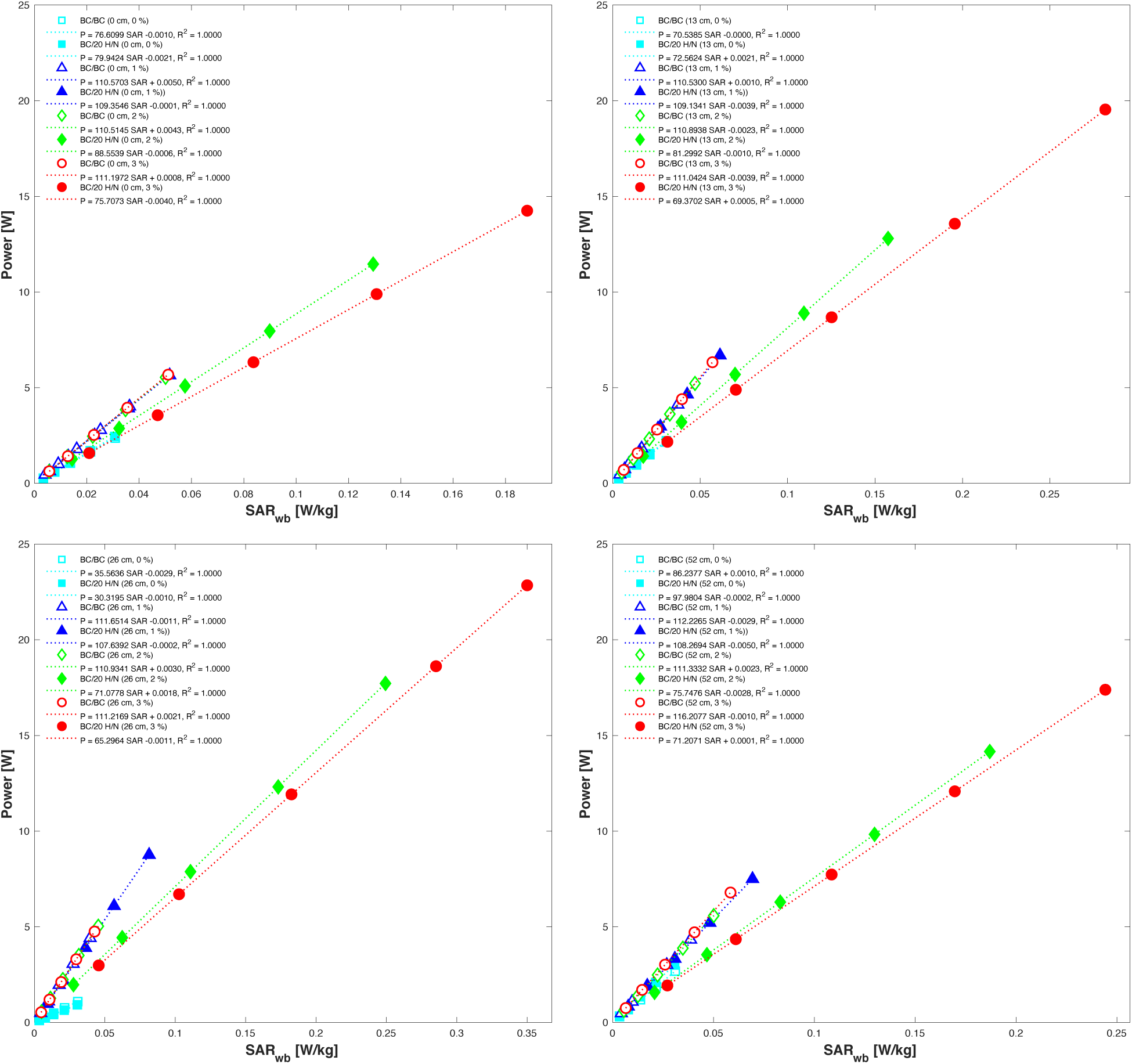
Plots of RF power vs. whole-body SAR for NaCl-H_2_O solutions with four conductive ionic concentrations, 0% (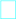), 1% (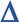), 2% (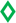) and 3% (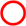), four conductive metallic implant wavelengths, 0, *λ*/2, *λ*, and 2*λ*, corresponding to four wavelengths, 0, 13, 26, and 52 cm, and two Tx/Rx combinations, BC/BC (open legends) and BC/20-H/N (solid legends) at 3T.

In **Figures 2** to **5** the plots with no conductive metallic implants (L = 0 cm, upper left plots) show the effects of the conductive ionic solutions (0, 1, 2, and 3% NaCl-H_2_O) and the plots with no conductive ionic solutions (0% NaCl-H_2_O, cyan lines and square legends 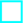) show the effects of the conductive metallic implant wavelengths of 0*λ*, *λ*/2, *λ* and 2*λ* (0, 26, 52, and 104 cm at 1.5T and 0, 13, 26, and 52 cm at 3T) on the RF power, B_1+rms_ and SAR_wb_ for RF coil Tx/Rx combinations of BC/BC (open legends) and BC/20-H/N (solid legends), at 1.5T and 3T, respectively.

Note that in **Figures 2** and **4**, corresponding to 1.5T, there were no differences in Tx/Rx combinations (solid legends were superimposed on the open legends), whereas in **Figures 3** and **5**, corresponding to 3T, there were differences in Tx/Rx combinations (solid legends were not superimposed on the open legends) in plots of P vs B_1_^2^ and P vs. SAR_wb_, respectively. This difference is due to the absence and presence of B_1_ shimming at 1.5T and 3T, respectively.

The R^2^ values of linear regression of B_1+rms_ vs. θ were less than 1 (R^2^<1) in **Table 1** because of the prescription of SB at 1.5T. The R^2^ values of linear regression were 1 for all fittings in **Table 2** because of the absence of SB at 3T, except for one, the regression of B_1+rms_ vs. θ (L=26 cm, C=3% and Tx/Rx = BC/20-H/N). The B_1+rms_ for θ = 90° was less than usual (0.7855 μT vs. 0.8525 μT), and a regression of B_1+rms_ of 0.7855 μT vs. θ for this experiment calculated a θ = 83° instead of 90°, which can be confirmed by the linearity in the plot of P vs. B_1_^2^ for this experiment (solid red legend) in **Figure 3**. The R^2^ values of linear regressions of P vs. B_1_^2^ and P vs. SAR_wb_ in **Tables 1** and **2** were equal to 1 (R^2^ = 1) at 1.5T and 3T, respectively.

The variation of RF power, B_1+rms_, whole-body specific absorption rate, transmit reference amplitude, transmit frequency, and absorption ratio were evaluated as a function of flip angle, RF coil transmit and receive combinations, conductive metallic wavelength, and ionic concentrations for 1.5T and 3T, respectively.

The boxplots of P (W), B_1+rms_ (μT), SAR_wb_ (W/kg), TxRA (V), TxF (MHz), and α vs. θ (30, 45, 60, 75, 90 degrees), Tx/Rx (1 and 2), L (0, 26, 52, and 104 cm at 1.5T, and 0, 13, 26, and 52 cm at 3T), and C (0, 1, 2, and 3%) are given in **Figs. 6** and **7** for 1.5T and 3T, respectively.

**Figure 6:**
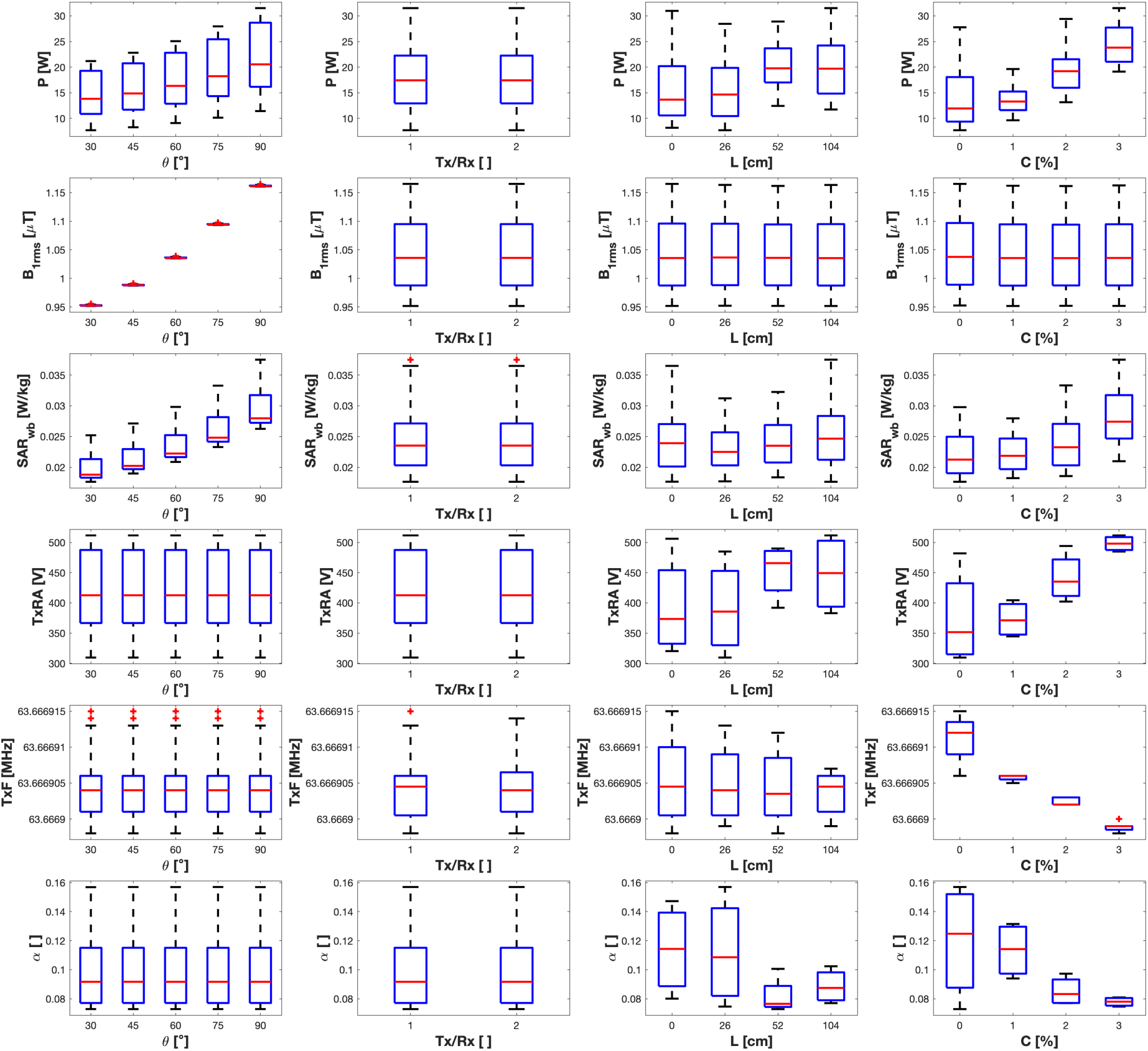
Boxplots of P (W), B_1+rms_ (μT), SAR_wb_ (W/kg), TxRA (V), TxF (MHz) and α vs. θ (30, 45, 60, 75, 90°), Tx/Rx (1: BC/BC, 2: BC/20-H/N), L (0, 26, 52, 104 cm) and C (0, 1, 2, 3%) at 1.5T.

**Figure 7:**
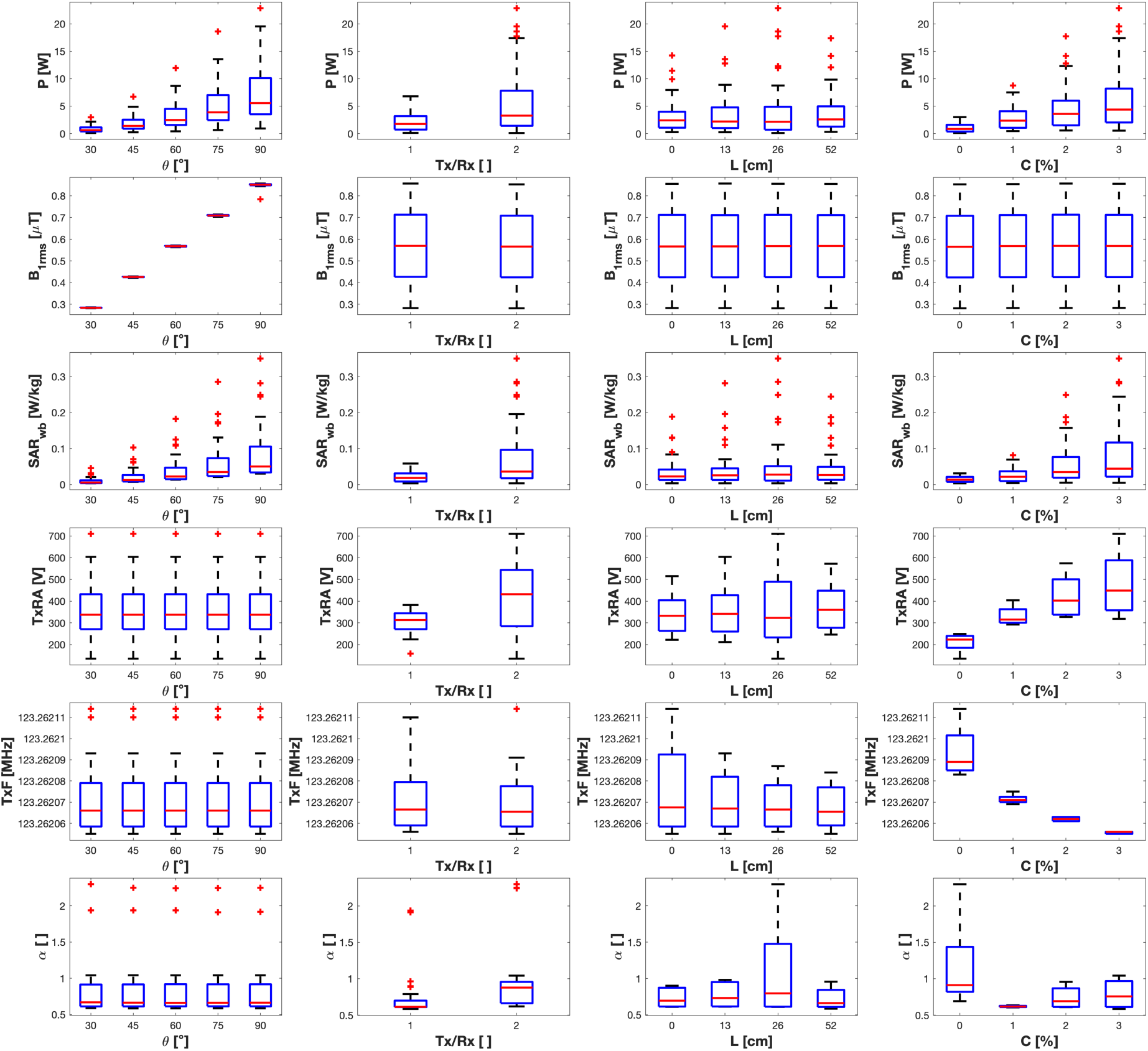
Boxplots of P (W), B_1+rms_ (μT), SAR_wb_ (W/kg), TxRA (V), TxF (MHz) and α vs. θ (30, 45, 60, 75, 90°), Tx/Rx (1: BC/BC, 2: BC/20-H/N), L (0, 13, 26, 52 cm) and C (0, 1, 2, 3%) at 3T.

The head and whole-body SAR scaled linearly with each other, depending on the Tx/Rx combinations for all experiments. The ratio, SAR_h_/SAR_wb_, was 7.95 and 7.95 at 1.5T, and 5.56 and 4.50 at 3T for Tx/Rx combinations of BC/BC and BC/H-20, respectively. There was no difference in Tx/Rx at 1.5T, but there was a difference at 3T. This was due to the differences in RF adjustments and B_1_ shimming at 1.5T and 3T, respectively.

P and B_1_ both increased with SB while maintaining the P/B_1_^2^ nearly constant, as in **Table 3**. The ratios of P and B_1+rms_ with and without SB, P_(SB)_/P and B_1(SB)_/B_1_, for five RF pulse flip angles (30, 45, 60, 75, 90 degrees) and equal sequence parameters (NS=3, T_R_=250ms, Tx/Rx=2) were 16.47, 7.84, 4.85, 3.47, 2.71 and 4.05, 2.80, 2.20, 1.86, 1.65 at 1.5T, and 11.31, 5.58, 3.56, 2.64, 2.31, and 3.36, 2.36, 1.89, 1.63, 1.46 at 3T, respectively. Note that the ratio of P_(SB)_/P scales with the square of the ratio of B_1(SB)_/B_1_ at both 1.5T and 3T.

P and B_1_^2^ varied linearly with NS and 1/T_R_ as given by Eq. (5). Both P and B_1_ increased with NS and decreased with T_R_ while maintaining a close ratio of P/B_1_^2^ when turning the SB on or off at both 1.5T and 3T as shown in **Table 3**. For example, the dP/dB_1_^2^ for five RF pulse flip angles and equal sequence parameters (NS=1, T_R_=250ms, Tx/Rx=2) with and without SB (1^st^ and 5^th^ rows) were 9.24 and 9.24 W/μT^2^ at 1.5T, and 7.77 and 7.82 W/μT^2^ at 3T, respectively.

As shown in **Table 3**, the B_1+rms_ varied linearly with θ per Eq. (1) independent of the B_0_ field when the same pulse sequence parameters were used. The variation of B_1+rms_ vs. θ was linear (R^2^=1) when SB was off, but nonlinear (R^2^<1) or quadratic (R^2^=1) when SB was on. The slope dB_1_/dθ and the intercept B_1_(0) varied distinctly when SB was on or off. The intercept corresponding to B_1+rms_ for zero-degree flip angle was zero, B_1_(0) = 0, when SB was off and nonzero, B_1_(0) ≠ 0, when SB was on. The slope, dB_1_/dθ, and intercept, B_1_(0), had a linear relationship when SB was on. The slope, dB_1_/dθ, decreased, and the intercept, B_1_(0), increased when SB was on.

P and B_1+rms_ increased with the thickness of SB in a step function while maintaining a close ratio of P/B_1_^2^ at 1.5T and 3T. The ratios of power and B_1+rms_ with and without SB, P_(SB)_/P and B_1(SB)_/B_1_, for three flip angles (30, 60, 90 degrees) were 5.02, 2.00, 1.44, and 2.23, 1.41, 1.20 for SB thicknesses of 3, 4, and 6 mm, and 11.32, 3.57, 2.14, and 3.36, 1.89, 1.46 for SB thicknesses of 12, 25, and 50 mm, respectively. The dP/dB_1_^2^ were 8.06, 8.04, and 8.06 W/μT^2^ for SB thickness of 0 mm, and SB thicknesses of 3, 4, and 6 mm, and SB thicknesses of 12, 25, and 50 mm, respectively.

B_1+rms_ vs. θ varied with SB thickness in a step function where dB_1_/dθ took one value, 0.0096 μT/deg, for SB thickness of 0 mm, and one value, 0.0066 μT/deg, for SB thicknesses of 3, 4, and 6 mm, and then another value, 0.0049 μT/deg, for SB thicknesses of 12, 25, and 50 mm.

The slice thickness (z), varied from 4 mm to 16 mm (4, 8 and 16 mm), did not have any effects on P, B_1+rms_, and SAR, and the linear regression coefficients, dB_1_/dθ, dP/dB_1_^2^, and dP/dS_wb_, for three flip angles (30, 60 and 90 degrees) at 1.5T and 3T.

## 4. DISCUSSION

In a previous study, the relationship between RF induced heating, RF power and B_1+rms_ was shown to vary with RF coils and Tx/Rx combinations, two Tx/Rx volume coils (a body coil and a head coil) and three Rx only phased-array coils (a 20-channel head and neck, a 32-channel head, and a 64-channel head and neck) in combination with a Tx/Rx body coil at one B_0_ field (3T) [35]. In this study, only the most widely used Tx/Rx combinations of RF coils (a body coil and a 20-channel head and neck coil) were used at two B_0_ fields (1.5T and 3T) to drastically reduce the number of experiments. Each additional RF coil would add 160 experiments for each B_0_ field and 320 experiments for both B_0_ fields.

Here, the relationship between the scanner-reported RF power, B_1+rms_, and SAR is evaluated as a function of RF pulse sequences, RF coil Tx/Rx combinations, conductive metallic wavelengths, and conductive ionic concentrations. The RF power and SAR varied with RF pulse sequences, RF coil Tx/Rx combinations, conductive metallic wavelengths, and conductive ionic concentrations, whereas B_1+rms_ varied with RF pulse sequences only, but not with Tx/Rx combinations, conductive metallic wavelengths, and conductive ionic concentrations.

As shown in **Tables 1** and **2**, the dB_1_/dθ did not change, whereas dP/dB_1_^2^ and dP/dS_w_ changed with the experimental parameters such as Tx/Rx, conductive metallic wavelengths, conductive ionic concentrations.

The RF transmit field (*B_1_*) is described by three components: the positive component (*B_1_*_+_), the negative component (*B_1_*_-_), and the magnitude of both positive and negative components (|*B_1_*|). The effective field that flips the spins in the rotating frame of reference in synchronization with the precession of the spins is the *B_1_*_+_ field, and it is defined as the time-averaged root-mean-square of the spatially averaged field (*B_1_*_+rms_) in the central axial slab [48]. The scanner adjusts the current and RF power in the transmit coil during the pre-scan mode until the desired *B_1_*_+_ field is reached for the prescribed flip angle [49–51]. As part of RF adjustments, the B_1_ shimming process adjusts the RF power and *E* field per RF coil Tx/Rx combinations while maintaining the B_1+_ field required for the prescribed flip angle [52–54]. This changes the relationship between the RF power and B_1+rms_ depending on the RF coils, Tx/Rx combinations, and experimental conditions [35]. As a result, RF power changes, but B_1_ remains nearly the same for the Tx/Rx combinations, conductive metallic wavelengths, and conductive ionic concentrations.

The dB_1_/dθ was equal, 0.0035 μT/deg at 1.5T, and varied slightly from 0.0094 to 0.0095 μT/deg with Tx/Rx at 3T for all of the experiments with varying conductive metallic wavelengths and conductive ionic concentrations as shown in **Tables 1** and **2** at 1.5T and 3T, respectively.

Initially, the differences in dB_1_/dθ were observed between the two B_0_ fields as 0.0035 μT/deg at 1.5T and 0.0095 μT/deg at 3T (**Tables 1** and **2**). This was further investigated and found that the difference was due to the differences in the prescription of SB and T_R_ in the nominal RF pulse sequences at 1.5T and 3T (**Table 3**). Note the effects of SB and T_R_ of 370 ms at 1.5T vs. no SB and the T_R_ of 250 ms at 3T, as shown in **Tables 1** to **3** at 1.5T and 3T.

When SB was on, the slope dB_1_/dθ decreased, but the intercept B_1_(0) increased at both 1.5T and 3T. Without SB, dB_1_/dθ was equal at both B_0_ fields (1.5T and 3T) at equal or scaled T_R,_ as seen in **Table 3**. The square of the ratios of dB_1_/dθ scaled with the ratios of 1/T_R_, for example, (0.0096/0.0079)^2^ equals (370/250) as shown in **Table 3**. P varied linearly with B_1_^2^ and 1/T_R_ per Eqs. (4) and (5) at both 1.5T and 3T. B_1+rms_ vs θ did not change with Tx/Rx combinations, but it changed with NS, T_R,_ and SB.

Since the RF pulse sequences with SB are not sensitive to the presence and absence of conductive metallic implants, conductive ionic solutions, implant wavelengths, or ionic concentrations, care must be taken to avoid SB because SB increases both P and B_1+rms_ drastically while maintaining the P/B_1_^2^ ratio nearly constant.

The dP/dB_1_^2^ varied from 8.4311 W/μT^2^ (Tx/Rx = 1 and 2, *λ*/2 = 26 cm, C = 0 %) to 23.3877 W/μT^2^ (Tx/Rx = 1 and 2, 2*λ* = 104 cm, C = 3 %) at 1.5T and from 1.2947 W/μT^2^ (Tx/Rx = 2, *λ* = 26 cm, C = 0 %) to 37.1368 W/μT^2^ (Tx/Rx = 2, *λ* = 26 cm, C = 3 %) at 3T with Tx/Rx, conductive metallic wavelengths and conductive ionic concentrations for all experiments (**Tables 1** and **2**). Since SAR is derived from RF power, their ratio can yield the absorption ratio as α = m·SAR_wb_/P per Eq. (3), which can reveal information about SAR computing algorithms of various scanners. A plot of P vs. SAR_wb_ gives a straight line with a slope of dP/dS_wb_, which equals the ratio of the patient’s weight to the absorption ratio (m/α) from which α can be measured per Eq. (3).

The dP/dS_wb_ varied from 433.1065 kg/α (Tx/Rx = 1 and 2, *λ*/2 = 26 cm, C = 0 %) to 933.1221 kg/α (Tx/Rx = 1 and 2, *λ* = 52 cm, C = 0 %) corresponding to α=0.157 to α=0.073 at 1.5T and from 30.3195 kg/α (Tx/Rx = 2, *λ* = 26 cm, C = 0 %) to 116.2077 kg/α (Tx/Rx = 1, 2*λ* = 52 cm, C = 3 %) corresponding to α=2.243 to α=0.585 at 3T, varying with Tx/Rx, conductive metallic wavelengths, and conductive ionic concentrations for all experiments (**Tables 1** and **2**).

The linear regression coefficients of P vs. SAR for head and whole-body, dP/dS_h_ and dP/S_wb_ scaled with each other linearly with ratios of 0.1257 and 0.1257 at 1.5T and ratios of 0.1798 and 0.2223 at 3T for Tx/Rx combinations of BC/BC and BC/20-H/N, respectively. The ratios of SAR_h_/SAR_wb_ were 7.9554 and 7.9554 at 1.5T and 5.5605 and 4.4977 at 3T for two RF coil Tx/Rx combinations. SAR_h_ and SAR_wb_ scaled by Tx/Rx for all experiments varying with conductive metallic implant wavelengths and conductive ionic concentrations.

The combination of body coil and head coil for transmit and receive had a significantly large effect on RF power, specific absorption rate, transmit reference amplitude, transmit frequency, and absorption ratio due to B_1_ shimming, which was available at 3T but not at 1.5T. The RF power and SAR for B_1+rms_ corresponding to a prescribed θ change significantly with the RF adjustment strategy and B_1_ shimming algorithms [47].

As shown in **Figures 6** and **7**, RF power and SAR changed with flip angles, Tx/Rx (3T), implant wavelength, and ionic concentration, whereas B_1+rms_ changed only with flip angles but not implant wavelength, and ionic concentration. The transmit reference amplitude, transmit frequency, and absorption ratio did not change with flip angle but changed with Tx/Rx (3T), implant wavelength, and ionic concentration.

Using the descriptive statistics, the mean plus/minus one standard deviation (m ± sd) for all experiments were 1.0467 ± 0.0750 μT and 0.5669 ± 0.2007 μT (B_1+rms_), 17.8827 ± 6.1112 W and 3.7476 ± 4.2007 W (RF power), 0.0241 ± 0.0045 W/kg and 0.0451 ± 0.0596 W/kg (SAR_wb_) at 1.5T and 3T, respectively.

These experiments show that the most important parameter is the RF power, as it varies significantly with the experimental conditions for a given B_1+rms_, which varies only with the RF pulse sequence but not the experimental conditions. RF power is most responsive to the presence or absence of conductive metallic implants and ionic solutions, metallic wavelengths, and ionic concentrations. While RF power and SAR both vary with the experimental conditions, the ratio between RF power and SAR is highly variable and unpredictable with the experimental conditions. Also, scanner-reported SAR computation for the RF pulse sequences may vary with the vendor-dependent algorithms and software versions.

RF power, B_1+rms_, and SAR relationship depends on the RF pulse sequences, RF coils, Tx/Rx combinations, RF adjustments and B_1_ shimming strategies, presence and absence of conductive metallic implants and conductive ionic solutions, implant wavelengths, and ionic concentrations. Experimental measurements are consistent with the theoretical formulations through the instrumentation proportionality constants, which can be determined for each of the RF coils and RF pulse sequences experimentally.

This experimental method provides means to simultaneously evaluate the relationship between the scanner-reported RF power, B_1+rms_, and SAR varying with RF pulse sequences, RF coils, Tx/Rx combinations, conductive metallic implant wavelengths, and conductive ionic concentrations during MRI. This experimental method can be used to evaluate specifically each of the MRI systems, RF coils, RF pulse sequences, conductive metallic implants, and conductive ionic solutions to optimize parameters for scanning patients with conductive metallic implants safely.

## 5. CONCLUSION

B_1+rms_ is insensitive to the presence and absence of conductive metallic implants and conductive ionic solutions, metallic wavelengths, ionic concentrations, RF coils, and Tx/Rx combinations. B_1+rms_ is not sufficient to monitor and control RF safety. B_1+rms_ varies with θ per Eq. (1) independent of B_0_ field, RF coils, Tx/Rx combinations, metallic wavelengths, and ionic concentrations using the same RF pulse sequences. The scanner-reported average RF power is responsive to all of the experimental conditions and remains the most reliable parameter to monitor and control RF safety. The scanner-reported RF power is sensitive to both metallic and ionic conductivities. The scanner-reported SAR scales with the scanner-reported average RF power linearly, with flip angles, but with different absorption ratios varying with experimental conditions, and remains unpredictable. Since the RF power was strongly associated with RF heating and temperature rise in a previous study, the response in RF power with experimental conditions must be considered for implant safety [35].

Prescribing RF saturation bands in the RF pulse sequences increases RF power, B_1+rms_, and SAR several orders of magnitude, especially when using low-power RF pulse sequences, and therefore saturation bands must be turned off when imaging patients with conductive metallic implants. The RF power, B_1+rms_, and SAR increase linearly with increasing number of slices and decrease with increasing repetition time.

## Data Availability

All data produced in the present work are contained in the manuscript

## ACKNOWLEDGEMENTS

This work was supported and performed at the Rockefeller Neuroscience Institute, Department of Neuroradiology, School of Medicine, West Virginia University, Morgantown, WV 26506 USA.

